# The Impact of COVID-19 on Mental Health outcomes among hospital fever clinic attendants across Nepal: A community-based cross-sectional study

**DOI:** 10.1101/2020.07.28.20163295

**Authors:** Hridaya Raj Devkota, Tula Ram Sijali, Ramji Bogati, Meraj Ahmad, Karuna Laxmi Shakya, Pratik Adhikary

**Affiliations:** Community Support Association of Nepal (COSAN), Kathmandu, Nepal; Manipal College of Medical Sciences, Pokhara, Nepal; Central Institute of Science and Technology (CIST) College (affiliated to Pokhara University), Nepal; UC Berkeley/Institute for Social and Environmental Research, Nepal (ISER-N)

**Keywords:** COVID-19, Depression, Anxiety, Stress, Pandemic, Public Health, Nepal

## Abstract

**Background:** The COVID-19 pandemic has been creating a panic and distressing situations among the entire population globally including Nepal. No study has been conducted assessing the psychological impact of this pandemic on the general public in Nepal. The objective of this study is to assess the mental health status during COVID-19 outbreak and explore the potential influencing factors among the population attending the hospital fever clinics with COVID–19 symptoms.

**Methods:** A cross-sectional survey was conducted between May - June, 2020 with a sample of 645 participants aged 18 and above in 26 hospitals across Nepal. Telephone interviews were conducted using a semi-structured questionnaire along with a validated psychometric tool, the Depression, Anxiety and Stress (DASS-21) scale. The metrics and scores of symptoms and their severity were created and analyzed. Multivariate logistic regression was used to determine the association of potential covariates with outcome variables.

**Results:** The prevalence of anxiety, depression and stress were 14%, 7% and 5% respectively. Participants from Bagmati province reported higher level of anxiety (OR 3.87, 95% CI 1.42 – 10.59), while stress (OR 4.78, 95% CI 1.09 – 21.29) and depressive symptoms (OR 3.37, 95% CI 1.10 – 10.35) observed higher among the participants in Province 1. Women were more at risk of anxiety (OR 4.26, 95% CI 2.21 – 8.20) and depression (2.75, 95% CI 1.16 – 6.51) than men. Similarly, people with primary level education found more prone to all factors, stress (OR 20.35, 95% CI 2.06 – 201.19), anxiety (OR 3.10, 95% CI 1.24 – 7.91), and depression (OR 4.41, 95% CI 1.29 – 15.07). More farmers than labors showed higher odds (OR 2.25, 95% CI 1.01 – 5.01) for anxiety, while individuals surveyed who reported their health status as poor-had higher odds (OR 5.95, 95% CI 1.08 – 32.68) for depression. Also, people currently living in rented houses reported more stress (OR 3.11, 95% CI 1.07 – 9.05) and those living far from family reported higher rates of depressive symptoms (OR 3.57, 95% CI 1.01 – 12.58).

**Conclusion:** The study identified increased prevalence of stress, anxiety and depressive symptoms during the initial stage of COVID-19 pandemic in Nepal. Considering the findings, there is urgent need to develop and implement appropriate community-based mental health programs targeting individuals who have had COVID-19 symptoms and who are prone to develop adverse mental health outcomes.

## Introduction

The World Health Organization (WHO) reports that as of June 26, 2020 (08:07 GMT), worldwide Covid- 19 has killed 492,085 with a total of 9,724,146 individuals confirmed infected (1), and the death toll is still rising. The scale and severity of the COVID-19 pandemic has threatened public health globally. The world has been reeling, even with high income countries in havoc as a result of the global spread of this potentially fatal disease. The WHO declared COVID-19, a Public Health Emergency on 30th January 2020 a month after the outbreak of the virus in Wuhan, China alerting the global community with particular concern to the high risk countries having poor health systems (1). Nepal in an example of a country that lacks adequate resources to tackle the COVID-19 outbreak. The government has not been able to assure the public that they are capable of handling the situation, and this has been created panic and distress throughout the entire population.

Nepal detected the first case of corona virus infection on 23rd January and the second case two months later, on 23rd March 2020 that surged to 11,700 affecting all 77 districts across the country with a total 28 reported deaths from COVID-19 by the end of June 2020 (2–4). The government strategy included a country-wide lockdown to prevent a widespread outbreak of the disease and this came into effect on 24th March 2020. This lockdown was partially lifted on 14th June 2020 (5). In addition to the illness itself, the entire population particularly the middle and low-income groups are already seriously affected through COVID-19 related issues such as lost jobs, restricted mobility and loss of freedom due to the nationwide lockdown as well as the on-going fear of disease susceptibility (6). Moreover, the government’s poor risk communication mechanisms and the strong influence of incorrect and misleading social media rumors has been creating further terror (7). All of these factors have negative mental health impacts on the public.

Previous studies that assessed the psychosocial impact of epidemics or pandemics such as SARS and COVID-19 found high levels of mental distress including panic attacks, and psychotic symptoms among healthcare workers and the general public (8–11). Evidence also shows that in addition to the stress of high numbers of people getting sick or dying, epidemics and pandemics also cause vast economic losses which are associated with further high psychosocial risk (12,13). It should also be noted that the most vulnerable groups – people who are poor, women, children, the elderly, persons with disability and the homeless, are reported to suffer during these public health emergencies and have the greatest difficulty rebuilding their means of subsistence and social support networks after such catastrophes (9,14). The effects on mental health are usually more marked among populations living under precarious circumstances, who have limited resources, and limited access to social support and healthcare services. The recent studies conducted in different settings during the COVID-19 pandemics reported comparable findings, with the highest levels of distress among women, rural inhabitants, elderly populations, groups with lower levels of education, migrant workers and those having unstable incomes (15,16). Moreover, perceived disease susceptibility and perceived disease severity (17), social isolation, and spending longer time watching COVID-19 related news and social media are also found associated risk factors with increased level of mental distress (18). WHO reports that the burden of distress, depression and other mental health conditions such as suicide is on the rise globally. It further reports that long-lasting moderate or severe depression related to COVID-19 pandemic may become a serious public health concern (19).

The burden of COVID-19 related mental disorders continues to grow with significant impacts on health and major social, human rights and economic consequences globally. Furthermore, mental health disorders, fear-related behaviors, stigmatization, and negative effect on access and quality of care during and in the aftermath of the epidemics are commonly reported (20,21). In summary, COVID-19 has the potential to create devastating social, economic and mental health crises which may have long-term impact, particularly in a country like Nepal.

In light of this, it is urgent to understand the current level of anxiety and distress due to COVID-19 in Nepal and to recommend evidence based mental health intervention policies to cope with this efficiently. It is further urgent that evidence-driven strategies be developed to reduce adverse psychological impacts and psychiatric symptoms during and after COVID-19. This study is intended to contribute to this effort by identifying potential factors that affect the mental health status of fever clinics patients with the symptoms of COVID–19. We hypothesized that the prevalence and levels of depression and anxiety as indicated on the respective Depression, Anxiety, and Stress Scale (DASS) subscales may be elevated among the study population.

## Methodology

### Study design and population

This study is a cross-sectional survey, conducted among the fever clinic attendants with symptoms of COVID-19 across various health care facilities in Nepal. The survey was conducted between May 17 to June 9, 2020 during the nationwide lock-down that started on 19^th^ March 2020.

### Participants’ recruitment procedure

The study covered all seven provinces across Nepal. A multi-stage sampling method was used for the selection and recruitment of participants. Twenty-six health facilities (hospitals and PHCs) from 23 districts were purposively selected ranging between 3 – 6 health facilities from each province covering both ecological zones, Hills and Terai. A sampling frame was developed collecting the names of those who attended hospital fever clinics between April 25 and May 16, 2020. Out of the total 1285 fever clinic attendants during the period, 687 met the study’s eligibility criteria and included in the final list for interviews. Individuals aged 18 and above, who visited hospital suspecting or having COVID-19 symptoms were inclusion criteria set for the study. The refusal rate for interviews was 6%. Figure 1 presents the number of participants approached for interviews and inclusion exclusion criteria used the study.

**Figure 1:**
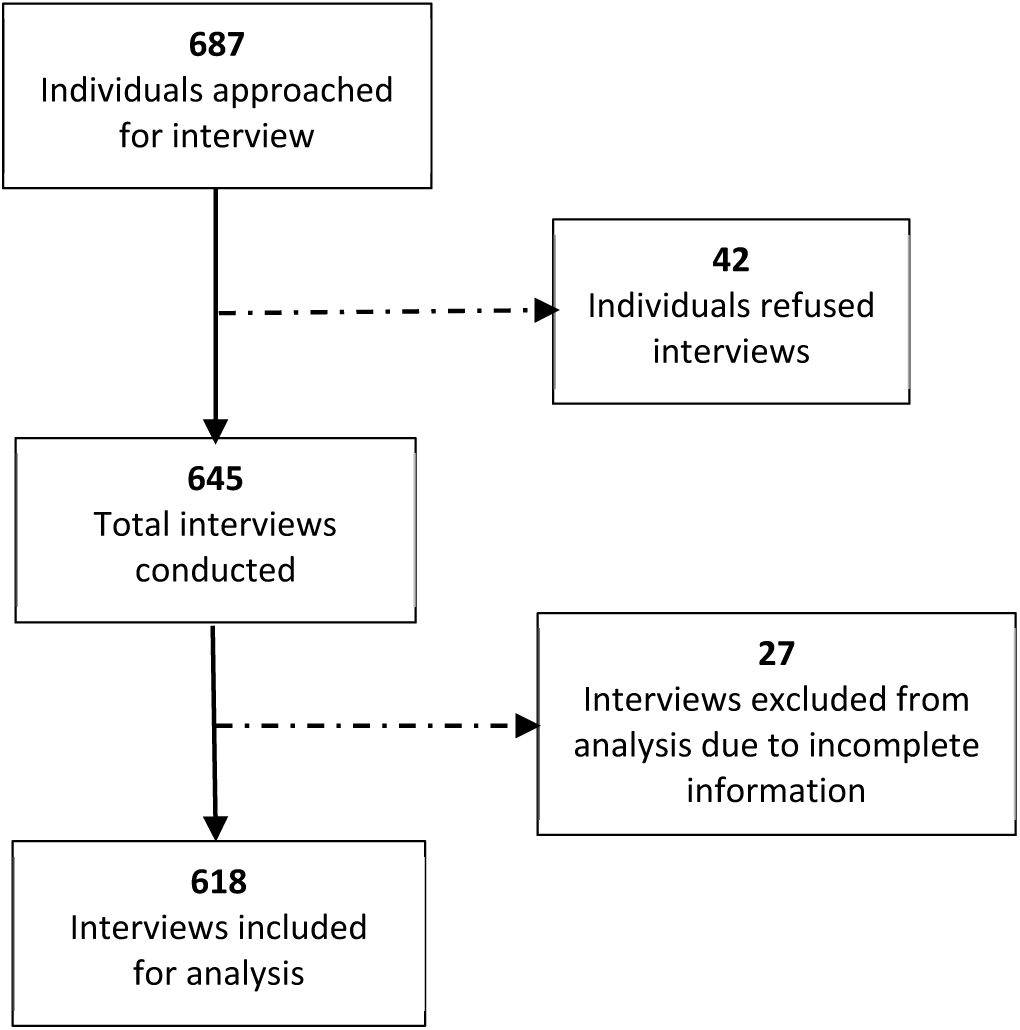
Flowchart of the sampling design and subject enrolment in the study

### Survey instrument and data collection procedure

A semi-structured questionnaire with socio-demographic information was used along with a 21- items depression, anxiety and stress scale (DASS-21) – a set of 3 self-reporting scales developed by Lovibond & Lovibond, 1995 (22) to measure the emotional states of depression, anxiety and stress were administered by the trained interviewers. The tool’s overall reliability coefficient (Cronbach’s Alpha) estimated 0.90. While estimating separately, it was 0.71, 0.79 and 0.77 for Depression, Anxiety and Stress respectively. The tool has already been tested in Nepal, its psychometric properties validated and it was found to be simple, easy to administer, and simple to score. It has been used extensively in previous studies globally as well as in Nepal (23–25).

Both the standardized questionnaire and DASS-21, were set up on tablet computers and mobile phones with KoBo Collect software, and administered in Nepali through telephone interviews by trained data collectors. The questionnaire was first developed in English, translated into Nepali by three bilingual Nepalese and field-tested for acceptability and comprehension among the population in which it was to be used. On average, administration of the questionnaire took 24 minutes.

### Ethical Approval

The researchers obtained ethical approval from the Nepal Health Research Council (NHRC) - ERB Protocol Registration No. 317/2020P. Before interviews, verbal informed consent was taken from all participants.

### Measures

DASS-21 score was the outcome variable that ranged from 0 – 42. The participant’s reaction to each statement was measured in a response category ranging from 0 – 3 to indicate “did not apply to me at all” to “applied to me very much most of the time”. Following the DASS – 21 scoring instructions, the sum of the rated scores in each statement was multiplied by 2 to calculate the final score (22). The cut- off (threshold) scores for detecting stress, anxiety and depression were 15, 8 and 10 respectively. The severity levels categorised as normal, mild, moderate and severe, and their score ranged as stated in the table below (Table 1).

**Table 1:** Severity level and score ranges

| Severity level | Stress | Anxiety | Depression |
| --- | --- | --- | --- |
| Normal | 0 - 14 | 0 - 7 | 0 - 9 |
| Mild | 15 - 18 | 8 - 9 | 10 - 13 |
| Moderate | 19 - 25 | 10 - 14 | 14 - 20 |
| Severe | 26 + | 15 + | 21 + |

**Table 2:** Variables and their definitions used in the study Note: The reliability coefficient (Cronbach’s Alpha) of 27-item knowledge questionnaire was estimated at 0.80.

| Variables | Definition |
| --- | --- |
| **Outcome Variables** |  |
| Stress, Anxiety and Depression | Total algebraic sum of the rated scores by the respondents (between 0 – 42) |
| **Covariates** |  |
| Province | Administrative division reported by the respondents as their permanent address. |
| Ecological region | The ecological (Hills or Terai) region of the respondent's habitation. |
| Place of residence | Respondent's place of residence – rural or urban at the time of survey. |
| Age | Completed age in years of respondent at the time of interview. |
| Gender | Self-reported sex identity of respondent - male or female) |
| Caste and ethnicity | Self-reported caste and ethnic group - Brahmin/Chhetri, Jana Jaati, Dalit, Madhesi, Muslims and others. |
| Religion | Self-reported religious belief of the respondent - Hinduism, Buddhism, Others. |
| Education | Number of years of education completed - no formal education, primary education, secondary education, higher education. |
| Primary occupation | Main occupation of the respondent at the time of interview - labor & others, service, foreign employment, farming, business and self-employment. |
| Marital status | Self-reported marital status of the respondent at the time of survey - married, ever married, single (widowed or divorced) |
| Family type | Husband, wife including the children considered a nuclear family, more than those living together is defined as joint or extended family. |
| Have own house | Respondent having his/her own house in any district or province - Yes or No |
| Current living | Respondent living (at his/her own house or rented house) at the time of survey |
| Living with family? | Respondent living with the family at the time of survey - Yes or No |
| Travel history | Recent travel outside the country - Yes or No |
| Health condition | Self-rated health status at the time of survey - very good, good, fair and poor. |
| Knowledge | Knowledge of symptoms, risk, transmission, and prevention spontaneously cited relating to COVID 19 presented as an additive score/index. It adapted 9 items related to symptom, 2 items risk, and 8 items each for transmission and prevention. |
| Perceived risk | An index developed using 6 questionnaire items at individual beliefs on safety, availability of health services, and government ability controlling the pandemic - nominal, medium, high. |

Socio-demographic data were self-reported by the participants. The knowledge and perceived risk indicators were created by 27 and 6 questionnaire items respectively to derive scores. The questionnaire included the questions related to COVID-19 symptoms, the risk groups, mode of transmission and prevention for knowledge assessment, while the individual’s belief about remaining safe from COVID-19, easy availability of healthcare services, and belief about government’s ability to control and overcome the pandemic was asked for risk perception. All the questionnaire items were equally weighted, dichotomized, and a composite measure was created using the sum with the maximum scores of 27 and 6 respectively.

### Statistical Analysis

The data collected in KoBo Collect software were downloaded into Excel Windows 10, cleaned and then transferred into SPSS (version 23.0 for Windows) for analysis. Descriptive statistics and associations between the outcome and potential covariates were examined using bivariate odds ratios (ORs) and 95% confidence intervals (CI). Due to the binary nature of the dependent variable, unconditional logistic regression was used. Any variable associated with the outcome variable with a p-value <0.2 in the bivariate analysis were further investigated for confounding by multivariate logistic regression model examining associations of potential covariates with outcome variables (stress, anxiety and depression).

## Results

### Characteristics of study participants

Out of 687 participants interviewed, a total of 618 were included in the analysis. Of these, the highest proportion of participants (17.8%) were from Karnali (Province 6) and the lowest from Province 5. The majority of study participants (79%) lived in urban area, while 63.6% in the hills. The average age of the participants was 35 years, with ages ranging from 18 – 85 (SD = 14.25). Over one-third (37%) were women, over half (55%) reported having secondary level education and 16% with higher education.

Nearly 4 in 10 reported their occupation as laborers and 16.8% reported having foreign employment. The vast majority of participants (86.1%) reported their religious belief as Hindu, with 42.71% and 17.8% reporting their caste group as Brahmin/Chhetri and Dalits respectively. More than 93% of respondents reported having their own house, however, only 83% were living in their house at the time of survey. About 77% of participants were married and 93% were living together with their family at the time of survey. (Table 4)

### Prevalence and severity measurement of stress, anxiety and depression

A considerable proportion of participants (14%) had symptoms of anxiety, while 7% and 5% reported depression and stress respectively. Over 5% had mild, 6% had moderate and just under 3% had severe levels of anxiety. Similarly, 3.4% had mild depression and 2.8% and 0.6% had the moderate and severe depressive symptoms. Under 3% had a mild level of stress and only 1.1% had moderate and another 1.3% had a severe level of stress. (Table 3)

**Table 3:** Prevalence and severity of stress, anxiety and depression

| Severity level (Score) | Stress |  | Anxiety |  | Depression |  |
| --- | --- | --- | --- | --- | --- | --- |
|  | n=618 | % | n=618 | % | n=618 | % |
| Normal (0 – 14) | 587 | 95.0% | 533 | 86.2% | 576 | 93.2% |
| Mild (15 – 18) | 16 | 2.6% | 33 | 5.3% | 21 | 3.4% |
| Moderate (19 – 25) | 7 | 1.1% | 37 | 6.0% | 17 | 2.8% |
| Severe (26 + ) | 8 | 1.3% | 15 | 2.4% | 4 | 0.6% |

Table 4 displays the result of the bi-variate logistic regression analysis. Among the participants across seven provinces, anxiety was found more prevalent in Bagmati (OR 3.79, 95% CI 1.60 – 8.98) and Sudurpaschim (OR 2.71, 95% CI 1.08 – 6.80). However, both stress and depression were reported as higher in Province 1 with OR 4.84, 95% CI 1.31 – 17.93 and OR 2.87 95% CI 1.12 – 7.37 respectively. Health condition, age, gender and marital status all showed the positive association with anxiety. The odds for those with poor health conditions (OR 3.72, 95% CI 1.08 – 12.83), the age group over 55 compared to the younger age (OR 2.86, 95% CI 1.35 – 6.07), women compared to men (OR 3.20, 95% CI 1.99 – 5.13) and divorced or widowed individuals compared to those with a partner (OR 5.00, 95% CI 1.80 – 13.88) were higher. Education also showed the positive association with both stress (P = 0.04) and anxiety (P = 0.02). Stress among the participants with primary and higher education level had higher odds (OR 8.82 95% CI 1.06 – 73.21) compared to those who did not have formal education. However, in the case of depression, participants with secondary level education had lower odds (OR 0.48, 95% CI 0.26 – 0.89). Individuals living in a rented house reported being more likely to develop both stress (OR 2.50, 95% CI 1.14 – 5.47) and anxiety (OR 1.78, 95% CI 1.03 – 3.07) than those living in their own house. Similarly, individuals living separately from their families reported having more stress compared to those living with their families (OR 2.70, 95% CI 0.98 – 7.42). Participants who recently travelled abroad were less likely to have anxiety compared to those who did not travel (OR 0.47, 95% CI 0.24 – 0.91).

### Factors associated with mental health outcome

Multivariate logistic regression analysis was performed to assess the relationship between outcome variables (stress, anxiety and depression) and independent variables that showed their relationship with a p-value <0.2 in the bivariate analysis (Table 4). After control of confounders, the province, and participants’ education showed significant association to all outcome variables. Those with primary and higher level education had higher odds than those with no school education for both stress and depression. The odds of those with primary education reported stress as OR 20.35, 95% CI2.06 – 201.19, and those from the higher education group was OR 14.63, 195% CI 1.21 – 176.5. It was OR 5.35, 95% CI 1.48 – 19.32 for those with primary and OR 6.04, 95% CI 1.24 – 29.56 for those with higher education in the case of depression. Only those with primary education were found to be associated to anxiety with higher odds (OR 3.10, 95% CI 1.21 – 7.91).

**Table 4:** Socio-demographic characteristics of the study participants with bivariate odds ratios (ORs)

| Covariate | Frequency (%) | Stress |  |  | Anxiety |  |  | Depression |  |  |
| --- | --- | --- | --- | --- | --- | --- | --- | --- | --- | --- |
|  | N=618 | OR | 95% CI | P-Value | OR | 95% CI | P-Value | OR | 95% CI | P-Value |
| **Province** |  |  |  |  |  |  |  |  |  |  |
| Karnali (6) | 110 (17.8%) | 1.00 |  |  | 1.00 |  |  | 1.00 |  |  |
| Gandaki (4) | 107 (17.3%) | 0.68 | 0.11 – 4.15 | 0.68 | 2.08 | 0.84 – 5.13 | 0.11 | 0.42 | 0.11 – 1.69 | 0.22 |
| Bagmati (3) | 96 (15.5%) | 3.69 | 0.97 – 14.05 | 0.06 | 3.79 | 1.60 – 8.98 | 0.002 | 1.34 | 0.47 – 3.84 | 0.59 |
| Province (1) | 92 (14.9%) | 4.84 | 1.31 – 17.93 | 0.02 | 1.56 | 0.59 – 4.12 | 0.37 | 2.87 | 1.12 – 7.37 | 0.03 |
| Sudurpaschhim (7) | 80 (12.9%) | 0.45 | 0.05 – 4.42 | 0.50 | 2.71 | 1.08 – 6.80 | 0.03 | 0.19 | 0.02 – 1.55 | 0.12 |
| Province (2) | 75 (12.1%) | 2.01 | 0.44 – 9.25 | 0.37 | 1.96 | 0.74 – 5.23 | 0.18 | 1.28 | 0.41 – 3.97 | 0.67 |
| Province (5) | 58 (9.4%) | 0.63 | 0.06 – 6.15 | 0.69 | 1.47 | 0.49 – 4.46 | 0.50 | 0.53 | 0.11 – 2.62 | 0.43 |
| **Ecological area** |  |  |  |  |  |  |  |  |  |  |
| Hills | 393 (63.6%) | 1.00 |  |  | 1.00 |  |  | 1.00 |  |  |
| Tarai | 225 (36.4%) | 1.11 | 0.53 – 2.33 | 0.79 | 0.79 | 0.48 – 1.29 | 0.34 | 1.20 | 0.64 – 2.28 | 0.57 |
| **Place of residence** |  |  |  |  |  |  |  |  |  |  |
| Urban | 488 (79%) | 1.00 |  |  | 1.00 |  |  | 1.00 |  |  |
| Rural | 130 (21%) | 0.54 | 0.19 – 1.58 | 0.26 | 1.37 | 0.81 – 2.33 | 0.24 | 0.74 | 0.32 – 1.70 | 0.47 |
| **Age** |  |  |  |  |  |  |  |  |  |  |
| 18 – 24 | 151 (24.4%) | 1.00 |  |  | 1.00 |  |  | 1.00 |  |  |
| 25 – 34 | 221 (35.8%) | 1.98 | 0.70 – 5.60 | 0.20 | 1.48 | 0.77 – 2.85 | 0.24 | 1.30 | 0.54 – 3.15 | 0.56 |
| 35 – 44 | 123 (19.9%) | 1.76 | 0.55 – 5.70 | 0.34 | 1.17 | 0.54 – 2.52 | 0.70 | 1.76 | 0.68 – 4.51 | 0.24 |
| 45 – 54 | 48 (7.8%) | 0.62 | 0.07 – 5.45 | 0.67 | 1.55 | 0.59 – 4.05 | 0.37 | 1.19 | 0.30 – 4.68 | 0.80 |
| 55 + | 75 (12.1%) | 1.65 | 0.43 – 6.32 | 0.47 | 2.86 | 1.35 – 6.07 | 0.01 | 1.28 | 0.40 – 4.05 | 0.68 |
| **Gender** |  |  |  |  |  |  |  |  |  |  |
| Male | 390 (63.1%) | 1.00 |  |  | 1.00 |  |  | 1.00 |  |  |
| Female | 288 (36.9%) | 1.44 | 0.69 – 2.97 | 0.33 | 3.20 | 1.99 – 5.13 | 0.000 | 1.78 | 0.95 – 3.34 | 0.07 |
| **Caste and Ethnicity** |  |  |  |  |  |  |  |  |  |  |
| Brahmin/Chhetri | 264 (42.7%) | 1.00 |  |  | 1.00 |  |  | 1.00 |  |  |
| Jana Jaati | 160 (25.9%) | 0.99 | 0.42 – 2.32 | 0.98 | 0.69 | 0.38 – 1.25 | 0.22 | 1.71 | 0.79 – 3.69 | 0.17 |
| Dalit | 110 (17.8%) | 0.15 | 0.02 – 1.17 | 0.07 | 0.86 | 0.45 – 1.63 | 0.64 | 1.03 | 0.39 – 2.75 | 0.95 |
| Madhesi/Muslims | 84 (13.6%) | 1.28 | 0.48 – 3.40 | 0.63 | 0.82 | 0.40 – 1.68 | 0.59 | 1.88 | 0.76 – 4.65 | 0.17 |
| **Religion** |  |  |  |  |  |  |  |  |  |  |
| Hinduism | 532 (86.1%) | 1.00 |  |  | 1.00 |  |  | 1.00 |  |  |
| Buddhism | 48 (7.8%) | 0.81 | 0.19 – 3.53 | 0.78 | 0.55 | 0.19 – 1.59 | 0.27 | 1.82 | 0.67 – 4.90 | 0.24 |
| Others | 38 (6.1%) | 1.04 | 0.24 – 4.55 | 0.96 | 1.14 | 0.46 – 2.83 | 0.77 | 2.37 | 0.87 – 6.48 | 0.09 |
| **Education** |  |  |  |  |  |  |  |  |  |  |
| No formal education | 98 (15.9%) | 1.00 |  |  | 1.00 |  |  | 1.00 |  |  |
| Primary (1 – 5) | 84 (13.6%) | 8.82 | 1.06 – 73.21 | 0.04 | 1.05 | 0.50 – 2.21 | 0.91 | 2.07 | 0.72 – 5.97 | 0.18 |
| Secondary (6 – 12) | 340 (55%) | 4.48 | 0.58 – 34.32 | 0.15 | 0.48 | 0.26 – 0.89 | 0.02 | 0.96 | 0.37 – 2.46 | 0.93 |
| Higher education | 96 (15.5%) | 8.82 | 1.08 – 71.92 | 0.04 | 1.03 | 0.50 – 2.12 | 0.95 | 1.02 | 0.32 – 3.29 | 0.97 |
| **Primary Occupation** |  |  |  |  |  |  |  |  |  |  |
| Labor | 240 (38.8%) | 1.00 |  |  | 1.00 |  |  | 1.00 |  |  |
| Service | 106 (17.2%) | 1.14 | 0.38 – 3.42 | 0.82 | 0.77 | 0.39 – 1.51 | 0.44 | 0.41 | 0.12 – 1.43 | 0.16 |
| Foreign employment | 104 (16.8%) | 1.41 | 0.50 – 3.98 | 0.52 | 0.58 | 0.28 – 1.22 | 0.15 | 1.49 | 0.65 – 3.40 | 0.35 |
| Farming | 94 (15.2%) | 1.02 | 0.31 – 3.34 | 0.97 | 1.21 | 0.64 – 2.28 | 0.55 | 1.13 | 0.45 – 2.83 | 0.80 |
| Business/Self-employed | 74 (12.0%) | 2.03 | 0.71 – 5.79 | 0.19 | 0.67 | 0.30 – 1.50 | 0.33 | 1.24 | 0.47 – 3.28 | 0.67 |
| **Marital Status** |  |  |  |  |  |  |  |  |  |  |
| Married | 475 (76.9%) | 1.00 |  |  | 1.00 |  |  | 1.00 |  |  |
| Ever Married | 127 (20.6%) | 1.15 | 0.48 – 2.74 | 0.76 | 0.80 | 0.43 – 1.47 | 0.47 | 0.52 | 0.20 – 1.34 | 0.18 |
| Single (Widow, Divorced) | 16 (2.6%) | 1.31 | 0.17 – 10.35 | 0.80 | 5.00 | 1.80 – 13.88 | 0.002 | 1.80 | 0.39 – 8.22 | 0.45 |
| **Family Type** |  |  |  |  |  |  |  |  |  |  |
| Extended | 451 (73.0%) | 1.00 |  |  | 1.00 |  |  | 1.00 |  |  |
| Nuclear | 167 (27.0%) | 1.11 | 0.50 – 2.46 | 0.80 | 0.93 | 0.56 – 1.57 | 0.80 | 0.96 | 0.47 – 1.95 | 0.90 |
| **Do you have own house?** |  |  |  |  |  |  |  |  |  |  |
| Yes | 576 (93.2%) | 1.00 |  |  | 1.00 |  |  | 1.00 |  |  |
| No | 42 (6.8%) | 0.94 | 0.22 – 4.10 | 0.94 | 1.79 | 0.83 – 3.90 | 0.14 | 0.67 | 0.16 – 2.87 | 0.59 |
| **Currently living @** |  |  |  |  |  |  |  |  |  |  |
| Own house | 514 (83.2%) | 1.00 |  |  | 1.00 |  |  | 1.00 |  |  |
| Rented | 104 (16.8%) | 2.50 | 1.14 – 5.47 | 0.02 | 1.78 | 1.03 – 3.07 | 0.04 | 1.18 | 0.53 – 2.62 | 0.69 |
| **Living with Family** |  |  |  |  |  |  |  |  |  |  |
| Yes | 574 (92.9%) | 1.00 |  |  | 1.00 |  |  | 1.00 |  |  |
| No | 44 (7.1%) | 2.70 | 0.98 – 7.42 | 0.05 | 1.96 | 0.93 – 4.13 | 0.08 | 2.36 | 0.94 – 5.95 | 0.07 |
| **Travel History** |  |  |  |  |  |  |  |  |  |  |
| No | 479 (77.5%) | 1.00 |  |  | 1.00 |  |  | 1.00 |  |  |
| Yes | 139 (22.5%) | 0.82 | 0.33 – 2.04 | 0.67 | 0.47 | 0.24 – 0.91 | 0.03 | 1.08 | 0.52 – 2.26 | 0.83 |
| **Knowledge (score)** |  |  |  |  |  |  |  |  |  |  |
| < 7 | 99 (16%) | 1.00 |  |  | 1.00 |  |  | 1.00 |  |  |
| 8 – 14 | 368 (59.5%) | 1.08 | 0.35 – 3.31 | 0.89 | 0.58 | 0.32 – 1.03 | 0.06 | 0.83 | 0.36 – 1.90 | 0.66 |
| 15 – 21 | 140 (22.7%) | 1.63 | 0.49 – 5.46 | 0.43 | 0.51 | 0.25 – 1.04 | 0.07 | 0.60 | 0.21 – 1.71 | 0.34 |
| 22 + | 11 (1.8%) | 5.28 | 0.85 – 32.90 | 0.08 | 0.88 | 0.18 – 4.39 | 0.87 | 2.53 | 0.47 – 13.76 | 0.28 |
| **Self-reported health** |  |  |  |  |  |  |  |  |  |  |
| Very good | 100 (16.2%) | 1.00 |  |  | 1.00 |  |  | 1.00 |  |  |
| Good | 333 (53.9%) | 0.54 | 0.21 – 1.39 | 0.20 | 0.69 | 0.35 – 1.37 | 0.29 | 0.80 | 0.33 – 1.97 | 0.63 |
| Fair | 171 (27.7%) | 0.74 | 0.27 – 2.05 | 0.56 | 1.79 | 0.90 – 3.56 | 0.10 | 1.09 | 0.42 – 2.84 | 0.86 |
| Poor | 14 (2.3%) | 2.21 | 0.41 – 11.91 | 0.35 | 3.72 | 1.08 – 12.83 | 0.04 | 3.62 | 0.82 – 16.08 | 0.09 |
| **Perceived Risk** |  |  |  |  |  |  |  |  |  |  |
| Nominal | 277 (44.8%) | 1.00 |  |  | 1.00 |  |  | 1.00 |  |  |
| Medium | 237 (38.3%) | 0.91 | 0.41 – 2.05 | 0.83 | 1.32 | 0.80 – 2.17 | 0.27 | 0.99 | 0.51 – 1.94 | 0.98 |
| High | 104 (16.8%) | 1.15 | 0.43 – 3.08 | 0.78 | 0.90 | 0.45 – 1.81 | 0.77 | 0.65 | 0.24 – 1.78 | 0.40 |

The adjusted odds for both stress (OR 4.78, 95% CI 1.09 – 21.03), and depression (OR 3.37, 95% CI 1.10 – 10.35) were higher among participants in province 1 than in Karnali. However, Bagmati had higher odds for anxiety (OR 3.87, 95% CI 1.42 – 10.59). Gender was associated to anxiety and depression with higher odds OR 4.26, 95% CI 2.21 – 8.20; and OR 2.75, 95% CI 1.16 – 6.51 respectively. Also, the current living status (owned or rented house) was significantly associated to stress, and living with or far from the family was associated to depression. The odds among those living in rented houses (OR 3.11, 95% CI 1.07 – 9.05) and far from their family (OR 3.57, 95% CI 1.01 – 12.58) were higher compared to those living in their own house and with family. Occupation and self-rated health conditions also showed their association to anxiety and depression respectively. The odds among farmers compared to labors, and those having poor health conditions than those with very good health were found to be higher. Their odds were calculated as OR 2.25, 95% CI 1.01 – 5.01 and OR 5.95, 95% CI 1.08 – 32.68 respectively. (Table 5)

**Table 5:**
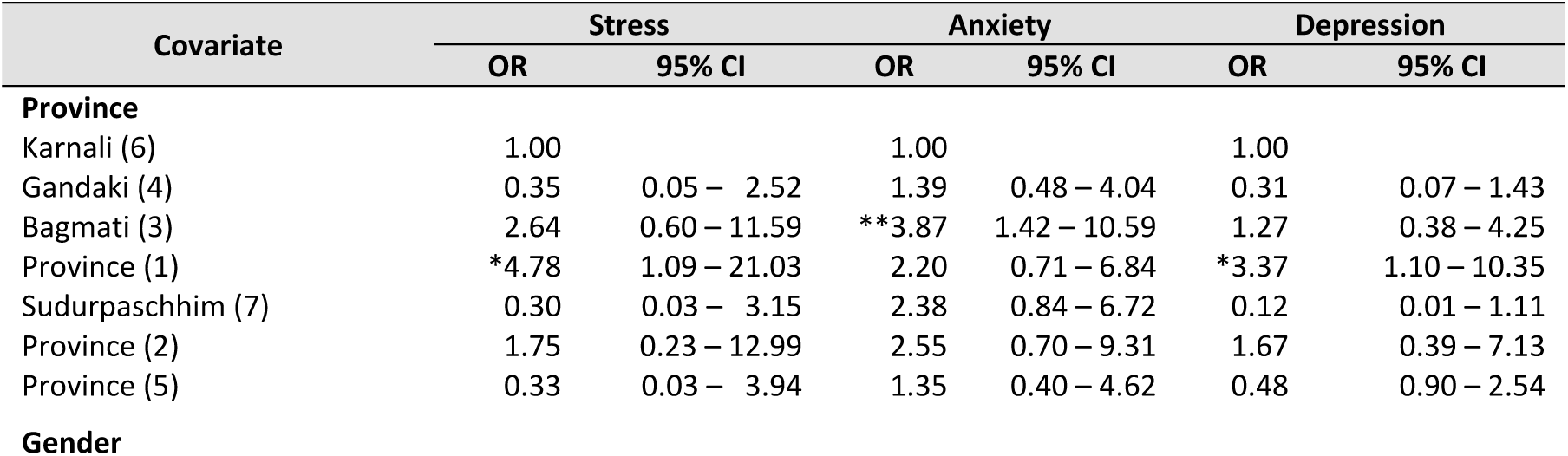
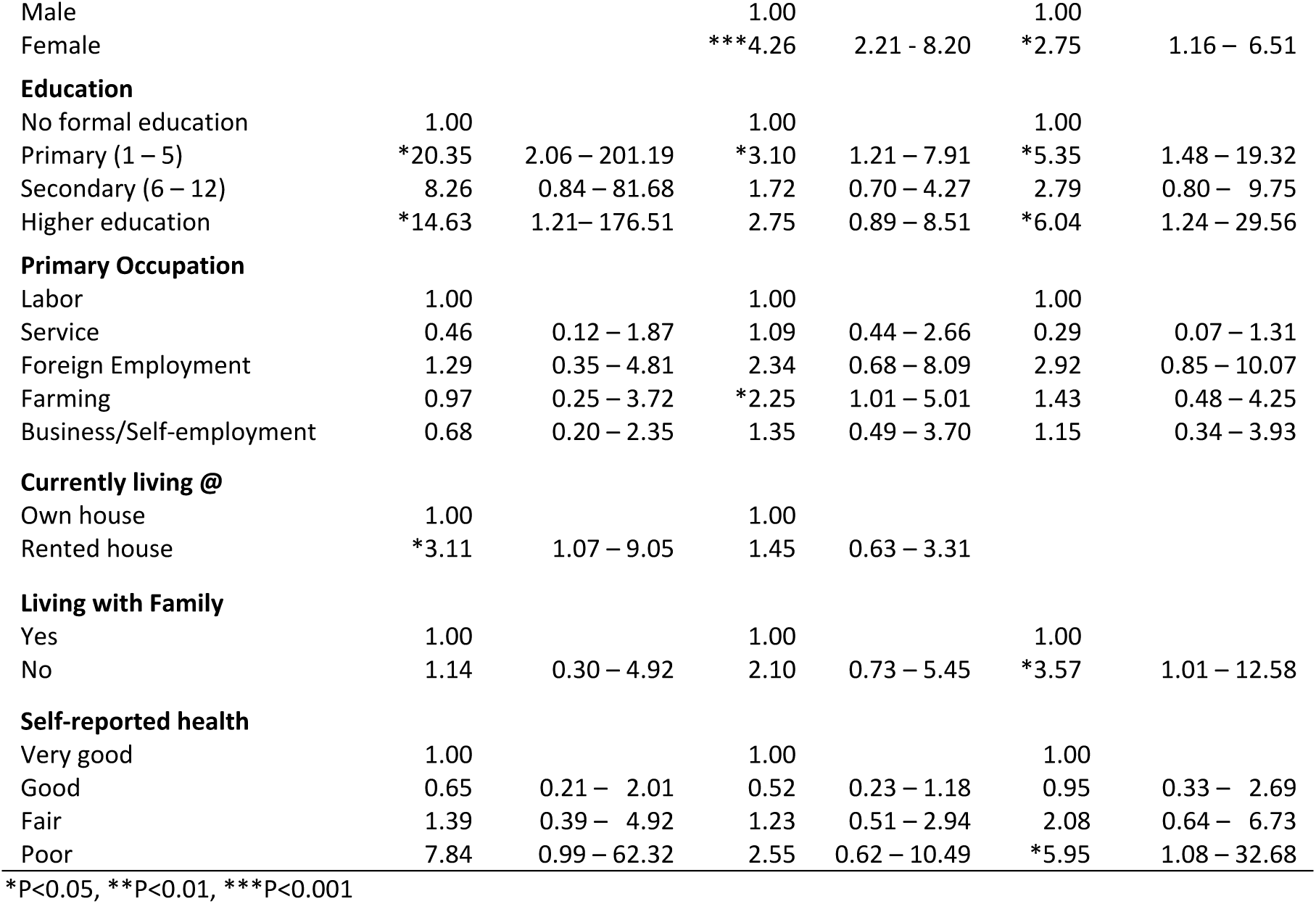
Multivariate analysis of stress, anxiety and depression by selected variables

| Covariate | Stress |  | Anxiety |  | Depression |  |
| --- | --- | --- | --- | --- | --- | --- |
|  | OR | 95% CI | OR | 95% CI | OR | 95% CI |
| **Province** |  |  |  |  |  |  |
| Karnali (6) | 1.00 |  | 1.00 |  | 1.00 |  |
| Gandaki (4) | 0.35 | 0.05 – 2.52 | 1.39 | 0.48 – 4.04 | 0.31 | 0.07 – 1.43 |
| Bagmati (3) | 2.64 | 0.60 – 11.59 | **\*\*3.87** | 1.42 – 10.59 | 1.27 | 0.38 – 4.25 |
| Province (1) | **\*4.78** | 1.09 – 21.03 | 2.20 | 0.71 – 6.84 | **\*3.37** | 1.10 – 10.35 |
| Sudurpaschhim (7) | 0.30 | 0.03 – 3.15 | 2.38 | 0.84 – 6.72 | 0.12 | 0.01 – 1.11 |
| Province (2) | 1.75 | 0.23 – 12.99 | 2.55 | 0.70 – 9.31 | 1.67 | 0.39 – 7.13 |
| Province (5) | 0.33 | 0.03 – 3.94 | 1.35 | 0.40 – 4.62 | 0.48 | 0.90 – 2.54 |
| **Gender** |  |  |  |  |  |  |
| Male |  |  | 1.00 |  | 1.00 |  |
| Female |  |  | ***4.26 | 2.21 – 8.20 | *2.75 | 1.16 – 6.51 |
| **Education** |  |  |  |  |  |  |
| No formal education | 1.00 |  | 1.00 |  | 1.00 |  |
| Primary (1 – 5) | *20.35 | 2.06 – 201.19 | *3.10 | 1.21 – 7.91 | *5.35 | 1.48 – 19.32 |
| Secondary (6 – 12) | 8.26 | 0.84 – 81.68 | 1.72 | 0.70 – 4.27 | 2.79 | 0.80 – 9.75 |
| Higher education | *14.63 | 1.21 – 176.51 | 2.75 | 0.89 – 8.51 | *6.04 | 1.24 – 29.56 |
| **Primary Occupation** |  |  |  |  |  |  |
| Labor | 1.00 |  | 1.00 |  | 1.00 |  |
| Service | 0.46 | 0.12 – 1.87 | 1.09 | 0.44 – 2.66 | 0.29 | 0.07 – 1.31 |
| Foreign Employment | 1.29 | 0.35 – 4.81 | 2.34 | 0.68 – 8.09 | 2.92 | 0.85 – 10.07 |
| Farming | 0.97 | 0.25 – 3.72 | *2.25 | 1.01 – 5.01 | 1.43 | 0.48 – 4.25 |
| Business/Self-employment | 0.68 | 0.20 – 2.35 | 1.35 | 0.49 – 3.70 | 1.15 | 0.34 – 3.93 |
| **Currently living @** |  |  |  |  |  |  |
| Own house | 1.00 |  | 1.00 |  |  |  |
| Rented house | *3.11 | 1.07 – 9.05 | 1.45 | 0.63 – 3.31 |  |  |
| **Living with Family** |  |  |  |  |  |  |
| Yes | 1.00 |  | 1.00 |  | 1.00 |  |
| No | 1.14 | 0.30 – 4.92 | 2.10 | 0.73 – 5.45 | *3.57 | 1.01 – 12.58 |
| **Self-reported health** |  |  |  |  |  |  |
| Very good | 1.00 |  | 1.00 |  | 1.00 |  |
| Good | 0.65 | 0.21 – 2.01 | 0.52 | 0.23 – 1.18 | 0.95 | 0.33 – 2.69 |
| Fair | 1.39 | 0.39 – 4.92 | 1.23 | 0.51 – 2.94 | 2.08 | 0.64 – 6.73 |
| Poor | 7.84 | 0.99 – 62.32 | 2.55 | 0.62 – 10.49 | *5.95 | 1.08 – 32.68 |

\*P<0.05, \*\*P<0.01, \*\*\*P<0.001

## Discussion

This study found 14% of the respondents with anxiety, 7% with depression and 5% with stress symptoms. Among them, moderate – severe level of anxiety was reported by 61%, and depression and stress by 50% and 48% respectively. The prevalence rates observed in the present study were not dramatically high compared to the recent studies conducted in other countries e.g. in China (18), and Italy (26) at the time of pandemics, however we found the prevalence of anxiety and depression higher than the estimated national prevalence rate (3.6% for anxiety and 3.2% for depression) at Nepal reported by WHO in 2017 (27). Our findings from this study were in line with the results of many other previous studies conducted in other countries. A study conducted during lockdown in India using the same tool (DASS-21) reported anxiety, depression and stress at 10%, 11% and 13% respectively (28). Moreover, a systematic review of COVID–19 and mental health literature conducted recently revealed the anxiety and depression prevalence between 16% – 28% and stress at 8% (9). Another study in Italy showed very high prevalence of depression at 32% and stress at 27% (26). The possible explanation for this wide variations in the prevalence could be the differences in the study populations, situations or the use of inconsistent definitions and instruments in the studies.

In the bivariate analysis, a number of environmental, socio-demographic and individual factors showed their relationship contributing to increased level of stress, anxiety and depression. For example, the living area (Province) was associated to all outcomes (stress, anxiety and depression), while the participant’s level of education and their stay at rented houses showed their association to increased stress and anxiety. The poorer people might be more likely to be renting a house who may have lost jobs due to lockdown, and that people with more education or less education may have felt threatened having different levels of understanding concerning COVID-19. Similarly, the participants living away from their family were found to be more likely to develop stress disorders. Moreover, age, gender, marital status, travel history and individual’s health condition with levels of elevated anxiety. After adjustment of potential confounders, only the participants’ living area (province), gender, education and self-reported health condition retained significant association with outcome variables. Also, the individual’s occupation and the variables related to living, in their own or in a rented home with or without family were found to be significantly associated to outcome variables in the multivariate regression model. Interestingly, the participant’s caste and ethnicity, knowledge about COVID-19 and their perceived risk did not show any relationship with mental health outcomes in this study.

The result of the multivariate regression analysis showed that the participants in province 1 were more prone to develop an increased level of stress and depression, while participants in Bagmati were found to be more likely to develop anxiety disorder compared to those in Karnali. There could be various explanations for this. A possible explanation may be that more information on the spread of the virus and its potential risks was available to people in Bagmati and Province 1 since these two provinces are more access to mass media compared to Karnali. Moreover, at the time of this study, the virus in Nepal was still perceived as being imported from other countries, the scope of community transmission to the rural province like Karnali was lower and the public in those area may have had yet to realize the pandemic’s scope in their territory.

Another finding from this study was that more women than men have been found to have increased levels of anxiety and depression during the outbreak of COVID-19. This finding is in line with previous studies conducted in China (10,15), India (28) and Italy (26), which have consistently found increased psychological distress higher among females compared to males. The finding may also be linked to evidence in the international literature that women likely to be more vulnerable to experiencing stress and developing post-traumatic symptoms. The explanation for this could be that women tend to feel increased responsibility to not only keep themselves well but maintain the health and well-being of their families – including older relatives, children and grandchildren that may have created more stress.

The present study also found an association between education and mental health outcome. The participants with formal education had higher prevalence of all mental health outcomes – stress, anxiety and depressive symptoms compared to those with not having school education. Previous studies in China (15) and Malaysia (29) identified similar findings - people with higher levels of education tended to have more distress. It could be because of high perceived risk of pandemics with self-awareness of their health, and also awareness of how it is spreading both nationally and internationally. Furthermore, they may have a better understanding of the short and long term impact – both in terms of health but also social and economic ramifications. The other factors, population with poor health condition, occupation as farmer, those living at rented house and far from the families found more at risk to develop depressive, anxiety and stress disorders. These people may also be poorer and at higher risk of immediate impact – with little savings or material goods they could sell if sick and unable to work, the impact of getting sick with COVID-19 could be immediate both for themselves and for members of their families. The finding, poor health condition showing association to adverse mental health outcome was consistently reported by the previous studies in China and Turkey.

The previous studies have been identified that the advanced age population, particularly those over 60 and comorbid with poor health were at most risk contracting the disease and also had a higher mortality to COVID – 19 (16,30). Corresponding to this, several studies conducted during this pandemic found the age and individual health condition highly linked to mental health outcome reflecting the increased psychological distress during the COVID-19. However, contrary to this, our study did not show any differences in increased level of stress, anxiety or depression between age groups. The possible reason for this could be the age cut-off value in this study at 55 years may have remained low to show this differences.

### Strength and limitation of the study

To the best of our knowledge, this is the first population study to evaluate the mental health status among Nepalese people during the COVID-19 pandemic. We feel this study is therefore, an important addition an important contribution to the literature as it provides preliminary data about the impact of COVID-19 on Napalese mental health. The strength of this study is a fairly large sample (n=618) recruited from 26 health facilities across the country representing main populations groups from different strata that included ecological region, urban rural residents and caste ethnicity. Moreover, the study had a high response rate (94%) that is considered good enough for telephone-based survey. Furthermore, the telephone interviews were taken by the well-trained and highly experienced interviewers under close supervision that ensured the quality of data collected.

With all these strengths, the study had a number of limitations. We intended to have a representative sample covering all province and ecological clusters, however, the purposive selection of health facilities conducting fever clinics may have resulted selection bias. Likewise, the potential sampling biases may have occurred with under or over representation from the different cluster, social groups, and also with the criteria as exclusion of under 18 populations and individual with communication and long-standing medical problems. Furthermore, the study population were only those who visited the fever clinic and recorded in the hospital register. However, a complete database of the clinic attending patients was not created across hospitals. Thus participants were selected purposively rather than random sampling procedure. It should also be noted that face to face interview was not possible due to lockdown and participants were interviewed over the phone on this sensitive issue. The absence of visual cues on the phone might have compromised creating comfortable environment for interview, rapport and probing (31). Also, the possibility of social desirability bias could not be ignored since the data were self-reported and there was no means of (clinical) verification.

## Conclusion

The result of this study shows an increased prevalence rate of mental health outcomes (stress, anxiety and depressive symptoms) among the study population. Interestingly however, the increased rates observed in the present study were not intensely high compared to the findings reported in previous studies conducted in the countries highly affected by COVID-19. This is noteworthy considering that this study was implemented in the initial phase of the pandemics in Nepal.

Most importantly, the study identified the key factors contributing to adverse mental health outcome and the population groups who are potentially more vulnerable to the pandemics. However, it is difficult to draw any conclusions regarding its long-term effect due to the cross-sectional nature of the current study. Further research is needed to track whether these groups show higher levels of psychosocial distress at later stages in pandemics. Longitudinal studies are recommended for understanding the trajectories of mental health among the population during and after the pandemic of COVID-19. Also, qualitative studies could be useful to understand how people cope with the pandemic and what psychosocial supports they have or feel they need in response to the pandemic.

One key policy implication of the present study is that the government should provide psychological support to all those already affected and who are prone to develop mental health concerns, not just the symptoms but the actual health problems need to be addressed during the pandemic. The individuals who are suffering through mental distress, and prone to develop serious symptoms in later stages must be reached with medical and counseling support. More attention needs to be given to vulnerable groups such as women, people with pre-existing illnesses and disabilities, farmers, and the migrant populations living away from their homes - far from their families and support networks.

## Data Availability

The manuscript itself present all the data used in this study. The data set can be available from the first author upon request.

## Declarations

The views expressed in this article are those of the authors and do not necessarily represent the views of supporting agency – Community Support Association of Nepal (COSAN).

We, the authors declare that we have no competing interest.

## Consent for publication

Not Applicable

## Availability of data and material

The datasets used for this study are available from the corresponding author upon request.

## Author’s contribution

HRD, PA and TRS conceived, designed and implemented the study in the field. MA, RB and KLS contributed to study design and undertook data collection mobilizing enumerators. HRD analysed data and wrote the manuscript with the help of TRS and PA. MA, RB and KLS provided their inputs for finalization of manuscript. All authors reviewed and approved the final version of the manuscript.

## Acknowledgement

The authors acknowledge the support and contribution of Community Support Association of Nepal (COSAN) and the colleagues who offered organizational facilities and staff time for this research. In addition, the authors wish to acknowledge the support provided by the hospitals and the data collectors – Shital Shrestha, Bhumika Sunuwar, Nabin Basnet, Asmita KC, Aayush Shrestha, Jeshika Shahi, Radhika Khadka, Dikshya Sharma, Sashi Bam, Ajay Poudel, Rakshya Adhikari, Aahana Sapkota, Prativa Pandey, Bibhushi Bhattarai, Saleena Shrestha, Aagya Dahal, Aaradhana Rayamajhi, Prashabdhi Shakya, Nisha Adhikari, Sapna Chaudhari, Deepa Ghimire and Nirmala Koju. The authors are grateful to those who participated in the study and shared their views and personal experiences.

## Author’s information

Hridaya Raj Devkota (PhD), Research Associate, Community Support Association of Nepal (COSAN), G.P.O. Box 6763, Bhagawati Marg, Kuleshwor-14, Kathmandu, Nepal Phone: 977(0) 1 4286781. (Corresponding author); Tula Ram Sijali (MA), Researcher, Community Support Association of Nepal (COSAN), Kathmandu, Nepal.; Ramji Bogati (PhD), Researcher, Community Support Association of Nepal (COSAN), Kathmandu, Nepal.; Meraj Ahmad (MPH), Asst. Professor, Manipal College of Medical Science, Pokhara, Nepal.; Karuna Laxmi Shakya (PhD), Central Institute of Science and Technology (CIST) College (affiliated to Pokhara University), Nepal.; Pratik Adhikary (PhD), Postdoctoral Researcher, UC Berkeley/Institute for Social and Environmental Research, Nepal (ISER-N).

## Reference

1. WHO. Coronavirus disease (COVID-19) Pandemic. 2020; Available from: https://www.who.int/emergencies/diseases/novel-coronavirus-2019

2. GoN/MoHP. Health Sector Response to Novel Coronavirus (2019-nCoV). SitRep #1 [Internet]. Kathmandu, Nepal; 2020. Available from: https://heoc.mohp.gov.np/update-on-novel-corona-virus-covid-19/

3. GoN/MoHP. Health Sector Response to Novel Coronavirus (2019-nCoV). SitRep # 43 [Internet]. Kathmandu, Nepal; 2020. Available from: https://heoc.mohp.gov.np/update-on-novel-corona-virus-covid-19/

4. GoN/MoHP. Health Sector Response to COVID-19. SitRep # 138 [Internet]. Kathmandu, Nepal; 2020. Available from: https://heoc.mohp.gov.np/update-on-novel-corona-virus-covid-19/

5. Kathmandu Post. “Fourth Nepali tests positive for Covid-19&”. kathmandupost.com. Archived from the original on 31 March 2020. Retrieved 27 June 2020. 2020; Available from: https://kathmandupost.com/national/2020/03/27/fourth-nepali-tests-positive-for-covid-19-case

6. Asmundson GJG, Taylor S. Coronaphobia: Fear and the 2019-nCoV outbreak. J Anxiety Disord. 2020;70(February).

7. The Himalayan Times. Youth arrested for spreading romours of COVID-19 cases”. The Himalayan Times. 2020 Mar 28; Available from: https://thehimalayantimes.com/kathmandu/youth-arrested-for-spreading-romours-of-covid-19-cases/

8. Maunder RG. Was SARS a mental health catastrophe? Gen Hosp Psychiatry [Internet]. 2009;31(4):316–7. Available from: http://dx.doi.org/10.1016/j.genhosppsych.2009.04.004

9. Ravi Philip Rajkumar. COVID-19 and mental health: A review of the existing literature. Asian J Psychiatr. 2020;52(January):102066.

10. Wang C, Pan R, Wan X, Tan Y, Xu L, Ho CS, et al. Immediate psychological responses and associated factors during the initial stage of the 2019 coronavirus disease (COVID-19) epidemic among the general population in China. Int J Environ Res Public Health. 2020;17(5).

11. Choi EPH, Hui BPH, Wan EYF. Depression and anxiety in Hong Kong during covid-19. Int J Environ Res Public Health. 2020;17(10).

12. Lai J, Ma S, Wang Y, Cai Z, Hu J, Wei N, et al. Factors Associated With Mental Health Outcomes Among Health Care Workers Exposed to Coronavirus Disease 2019. JAMA Netw - Open. 2020;3(3):1–12.

13. PAHO/WHO. PROTECTING MENTAL HEALTH DURING EPIDEMICS. 2006;1:1–25.

14. Tsai J, Wilson M. COVID-19: a potential public health problem for homeless populations. Lancet Public Heal [Internet]. 2020;5(4):e186.#x2013;7. Available from: http://dx.doi.org/10.1016/S2468-2667(20)30053-0

15. Qiu J, Shen B, Zhao M, Wang Z, Xie B, Xu Y. A nationwide survey of psychological distress among Chinese people in the COVID-19 epidemic: Implications and policy recommendations. Gen Psychiatry. 2020;33(2):19–21.

16. Wang B, Li R, Lu Z, Huang Y. Does comorbidity increase the risk of patients with covid-19: Evidence from meta-analysis. Aging (Albany NY). 2020;12(7):6049–57.

17. Gu J, Zhong Y, Hao Y, Zhou D, Tsui H, Hao C, et al. Preventive behaviors and mental distress in response to H1N1 among university students in Guangzhou, China. Asia-Pacific J Public Heal. 2015;27(2):pNP1867–79.

18. Gao J, Zheng P, Jia Y, Chen H, Mao Y, Chen S, et al. Mental health problems and social media exposure during COVID-19 outbreak. PLoS One [Internet]. 2020;15(4):1–10. Available from: http://dx.doi.org/10.1371/journal.pone.0231924

19. World Health Organization. Mental disorder: Key Facts [Internet]. 28 November. 2019 [cited 2020 Jun 27]. Available from: https://www.who.int/en/news-room/fact-sheets/detail/mental-disorders

20. Leary AO, Jalloh MF, Neria Y. Fear and culture?: contextualising mental health impact of the 2014 – 2016 Ebola epidemic in West Africa. 2018;(August 2014):1–5.

21. Zhong B, Luo W, Li H, Zhang Q, Liu X, Li W, et al. Knowledge, attitudes, and practices towards COVID-19 among Chinese residents during the rapid rise period of the COVID-19 outbreak?: a quick online cross-sectional survey. Int J Biol Sci. 2020;16:1745–52.

22. Lovibond S &, Lovibond P. Manual for the depression, anxiety, and stress scales (2nd ed). Sydney, New South Wales, Australia: Psychology Foundation. 1995;

23. Lee EH, Moon SH, Cho MS, Park ES, Kim SY, Han JS, et al. The 21-Item and 12-Item Versions of the Depression Anxiety Stress Scales: Psychometric Evaluation in a Korean Population. Asian Nurs Res (Korean Soc Nurs Sci) [Internet]. 2019;13(1):30–7. Available from:https://doi.org/10.1016/j.anr.2018.11.006

24. Lumley M, Katsikitis M, Statham D. Depression, Anxiety, and Acculturative Stress Among Resettled Bhutanese Refugees in Australia. J Cross Cult Psychol. 2018;49(8):1269–82.

25. Tonsing KN. Psychometric properties and validation of Nepali version of the Depression Anxiety Stress Scales (DASS-21). Asian J Psychiatr [Internet]. 2014;8(1):63–6. Available from: http://dx.doi.org/10.1016/j.ajp.2013.11.001

26. Mazza C, Ricci E, Biondi S, Colasanti M, Ferracuti S, Napoli C, et al. A nationwide survey of psychological distress among italian people during the covid-19 pandemic: Immediate psychological responses and associated factors. Int J Environ Res Public Health. 2020;17(9):1–14.

27. World Health Organization. Depression and Other Common Mental Disorders: Global Health Estimates. Geneva; 2017.

28. Rehman U, Shahnawaz MG, Khan NH, Kharshiing KD, Khursheed M, Gupta K, et al. Depression, Anxiety and Stress Among Indians in Times of Covid-19 Lockdown. Community Ment Health J [Internet]. 2020; Available from: http://www.ncbi.nlm.nih.gov/pubmed/32577997

29. Lee KW, Ching SM, Hoo FK, Ramachandran V, Chong SC, Tusimin M, et al. Prevalence and factors associated with depressive, anxiety and stress symptoms among women with gestational diabetes mellitus in tertiary care centres in Malaysia: A cross-sectional study. BMC Pregnancy Childbirth. 2019;19(1):1–11.

30. Zhou F, Yu T, Du R, Fan G, Liu Y, Liu Z, et al. Clinical course and risk factors for mortality of adult inpatients with COVID-19 in Wuhan, China: a retrospective cohort study. Lancet [Internet]. 2020;395(10229):1054–62. Available from: http://dx.doi.org/10.1016/S0140-6736(20)30566-3

31. Novick G. Is there a bias against telephone interviews in qualitative research? Res Nurs Heal. 2008;31(4):391–8.

